# Predictors of Road Safety behaviors among Boda-Boda Operators and their passengers in Kampala: A Mixed-Methods Study

**DOI:** 10.64898/2026.05.29.26354085

**Authors:** Rose Ainembabazi, Derrick Kimuli, Taibu Murami, Solomon T. Wafula, Evance Mgeyi, John Bosco Kwesiga, Pascal Kibingo, Ivan Mugumya, Nnodimele Onuigbo Atulomah, Deo Nsubuga

**Affiliations:** Faculty of Science and Technology, Department of Health Sciences, Cavendish University Uganda, Kampala, Uganda; Ministry of Health, Department of Public Health, Kampala, Uganda; Makerere University School of Public Health, Kampala, Uganda; Department of Programs, Baylor College of Medicine Children’s Foundation, Mwanza, Tanzania Babcock University

**Keywords:** Boda-boda safety, Road traffic injuries, Safe riding behaviors, Occupational road safety

## Abstract

1.

**Background:** Despite road safety regulations, “Boda Boda” riders in Uganda continue experiencing frequent road traffic injuries due to unsafe riding behaviours, increasing morbidity and mortality among riders and passengers. Evidence on safe riding practices and associated factors remains limited. This study assessed factors associated with safe riding behaviours among boda-boda riders and passengers in Kampala Central Division.

**Methods:** A cross-sectional survey using a convergent parallel mixed-methods design guided by Predisposing, Reinforcing, and Enabling Constructs in Educational/Environmental Diagnosis and Evaluation Model was conducted. Quantitative data were collected from 424 riders using structured questionnaires and analyzed using descriptive statistics and binary logistic regression to determine predictors of safe riding behaviours, with adjusted odds ratios (AORs) reported at p<0.05. Qualitative data were collected simultaneously through in-depth semi-structured interviews with 10 passengers. Thematic analysis and triangulation were used to identify convergences and divergences, providing a comprehensive understanding of safety determinants among riders and passengers.

**Results:** Among 424 riders (mean age 29.56 ± 5.71), 65.1% exhibited unsafe riding behaviors. Bivariate analysis identified education, marital status, religion, willingness to obey traffic regulations, and family encouragement as significant factors. In the adjusted model, secondary (AOR=0.50; 95% CI:0.30–0.85) and post-secondary education (AOR=0.57; 95% CI:0.33–0.98), being married (AOR=0.56; 95% CI:0.34–0.91), Christian religion (AOR=2.98; 95% CI:1.63–5.47), willingness to obey traffic regulations (AOR=0.41; 95% CI:0.24–0.70), union advocacy (AOR=1.76; 95% CI:1.03–3.01), and well-maintained roads (AOR=1.65; 95% CI:1.07–2.55) were significant predictors. Qualitative findings highlighted barriers including helmet lack, over-speeding, traffic regulation disregard, and poor road infrastructure

**Conclusions:** Rider and passenger safety is still low, interdependent, and influenced by multiple factors. Integrated interventions focusing on education, stronger families, religious affiliations, union safety advocacy, and stricter enforcement of traffic regulations are vital for enhancing safety for both riders and passengers.

Clinical trial number: not applicable

## 2. Background

Injuries from Road traffic accidents remain one of the leading public health concerns globally, accounting for an estimated 1.35 million deaths or disabilities every year^1^. While there has been a decline in global fatality rates per 100,000 people since 2010, the burden remains disproportionately higher in low- and middle-income countries (citation required at least two). In sub-Saharan Africa, studies highlight the severe consequences of injuries from road traffic accidents, with head injuries accounting for a large share of admissions and mortality^2 3 4^.

In Uganda, although RTIs significantly declined from 22,461 to 13,244 crashes between 2009 and 2017, severe RTIs increased from 14.7% to 22.2%, with motorcycle (boda-boda) involvement rising from 24.5% to 33.9% in the same period^5^. Police reports indicate that boda-boda were responsible for over 40% of road traffic fatalities in 2019^6^. Hospital-based studies also indicated that RTIs contribute up to 40% of surgical cases at Mulago National Referral Hospital^7 8^. In addition, the government of Uganda incurs treatment costs estimated between USD 245–3590 per accident victim^9^, straining already limited healthcare resources.

Previous studies attribute increased rates of accidents to human error, risky riding practices, poor road infrastructure, and limited enforcement of traffic regulations^10,11^. Head injuries, over-speeding, low helmet use, and inadequate trauma care compound the burden^12,13^. While initiatives such as “Safe-Boda” have introduced structured training and promoted helmet use among motorcycle riders and their clients^14,15^, these interventions remain limited in reach and adoption. Extant research has further highlighted defects in rider training, poor compliance with licensing and traffic rules, weak law enforcement, and a lack of consistent safety gear use^16–20^.

Despite the growing body of literature on road safety behaviors, studies specifically focusing on the factors associated with both passenger and rider safety in the boda-boda sector remain limited. Existing work often emphasizes general road traffic safety rather than the unique occupational risks of boda-boda riders, whose workplace is the road itself^20,21^. This gap in evidence limits the design and deployment of targeted and sustainable interventions. This study, therefore, applies the PRECEDE model to examine predisposing, reinforcing, and enabling factors associated with passenger and rider safety in Kampala city, Uganda. By integrating quantitative and qualitative approaches, it contributes context-specific evidence to inform policy, enforcement, training, and infrastructure interventions that can reduce boda-boda–related accidents and improve workplace safety for riders and their passengers.

## 3. Methods

### 3.1. Study design and participants

A cross-sectional, convergent parallel mixed-methods design was used, guided by the PRECEDE model. Quantitative and qualitative data were collected simultaneously and independently. The study was conducted in Kampala Central Division, an area characterized by intense traffic and high boda-boda activity.

**Riders (quantitative):** Boda-boda riders operating in Kampala Central Division, aged ≥18 years, with ≥6 months’ riding experience, and willing to participate were recruited from 05/02/2026-28/02/2026. Riders with <6 months’ experience, those <18 years, or unwilling to participate were excluded.

**Passengers (qualitative):** For the qualitative component, the grounded theory framework with etic perspectives was applied to understand the barriers to practicing safe riding behaviors. Multi-variation Purposive sampling was used to select passengers aged ≥18 who had used boda-boda services regularly for the previous 6 weeks; irregular users and those who opted out were excluded.

### 3.2. Sampling and Sample Size

Sampling frame and selection: Boda-boda stages (designated pick-up/drop-off points) in Kampala Central Division were enumerated. 106 stages were selected by simple random sampling. From each selected stage, riders were chosen by systematic sampling (every 4th rider) to achieve the target sample and reduce clustering. Sample size calculation (riders): For an unknown population size, the sample size computational formula for single-proportion used in a study by Leslie Kish (1965) was used with 95% confidence level on the standard normal distribution curve, P=0.5 (for a conservative estimate; due to no studies on road safety behavior index), a maximum tolerable sampling error of 0.05 and a 10% for non-response to yield a final target of 424 riders. Qualitative sample (passengers) was guided by data saturation, which was defined as the point at which no new themes were yielded during in-depth interviews. At the end,10 in-depth interviews were conducted^22 23 24^

### 3.3. Data Sources and Instruments

Structured rider questionnaire was developed consisted of (demographics; predisposing factors of knowledge [2-items on a binary response pattern of Yes/No], attitudinal dispositions [3-items on a agree/disagree response pattern], perceptions [4-items on a agree/disagree response pattern]; reinforcing [5-items on a agree/disagree response pattern]; enabling [3-items on a agree/disagree response pattern]; and riding behaviors (11-items transformed to 2 items of safe and unsafe riding behaviour). Items in the instrument were closed-ended and Likert-type responses. Semi-structured passenger interview guide addressing experiences, perceived effectiveness of safety measures, and stakeholder roles. Public Health Experts, Government reports (e.g., Uganda Police; Ministry of Works and Transport), and published literature on road safety and boda-boda operations informed context and instrument development.

### 3.4. Procedures

Trained research assistants administered boda-boda rider questionnaires at sampled stages. Passenger qualitative interviews were conducted using an interview guide with open-ended questions. Quantitative and qualitative strands were implemented in parallel and analyzed independently before integration.

### 3.5. Study variables and measurement

The outcome variable was self-reported riding behavior and measured using eleven (11) items assessed on a four-frequency response Likert-type scale: none of the time, some of the time, most of the time, and all the time. Responses were recoded from 0 to 3, where 3 represented the most desirable riding behavior, and 0 represented the least desirable behavior. Scores from the eleven items were summed to generate an aggregate riding behavior score for each respondent.

Normality of the aggregated riding behavior score was assessed using the Shapiro–Wilk test. The test indicated that the data significantly deviated from a normal distribution (p < 0.05). Log transformation was attempted, but the distribution remained non-normal. Consequently, the riding behavior variable was dichotomized using the median score. Respondents with scores above the median were categorized as practicing safe riding behaviors, whereas those with scores equal to or below the median were categorized as practicing unsafe riding behaviors.

The independent variables included socio-demographic characteristics such as age, education level, marital status, and religion. Additional explanatory variables were grouped into domains based on PRECEDE model constructs, including predisposing factors, reinforcing factors, and enabling factors.

Predisposing factors included knowledge, attitudes, and perceptions related to road safety. Knowledge was measured using two items assessing whether the rider knew that helmet use is important and that over-speeding can cause accidents, with response options coded as Yes or No. Attitudes were assessed using statements such as willingness to obey traffic regulations, willingness to ride with caution, and fear of accidents when riding recklessly. Perceptions included beliefs that helmets protect the head during crashes, motorcycle accidents can be serious, anyone can be involved in an accident, and reckless riders are more likely to be involved in accidents.

Reinforcing factors included family encouragement of road safety practices, adherence to safety rules by other riders, advocacy for safety by boda-boda associations, government safety programs that increase road safety awareness, and access to affordable helmets and reflective jackets. Enabling factors included road conditions used by boda-boda riders, adequacy of road safety training received, and the availability of safety measures on the roads.

Except for the knowledge items, the explanatory variables were measured using a four-point Likert scale consisting of Strongly Agree (SA), Agree (A), Disagree (D), and Strongly Disagree (SD). Responses were coded on a scale of 0 to 3, where 3 represented the most desirable response, and 0 represented the least desirable response. Due to low frequencies in some response categories, the variables were recoded into binary categories, where scores of 0, 1, and 2 represented disagreements, while a score of 3 represented agreement. These variables were therefore included in the analysis as binary variables.

To ensure validity, the questionnaire was reviewed by subject matter experts in public health and road safety to determine whether the items adequately captured the intended constructs. Necessary revisions were made based on their recommendations to improve clarity and content coverage.

### 3.6. Data Analysis

Quantitative data were analyzed using IBM SPSS Statistics version 22. Descriptive statistics were used to summarize the socio-demographic characteristics of the riders and the scores of the different components of the PRECEDE model. Binary logistic regression analysis was conducted to examine the association between predictor variables (predisposing, reinforcing, and enabling factors) and the boda-boda riding Behaviors.

At the bivariate level, each independent variable was analyzed separately against the outcome variable using a chi-square test of independence for categorical variables, while an independent t-test was used to test the association with age (continuous variable). Variables with a p-value ≤ 0.25 were considered eligible and entered the multivariable binary logistic regression model using the enter method to control for potential confounding. Adjusted Odds Ratios (AORs) with 95% Confidence Intervals (CIs) were reported to determine the strength of association. Statistical significance in the final model was set at p < 0.05.

For the qualitative component, interview transcripts were analyzed using Thematic Analysis. The transcripts were reviewed and coded to identify recurring patterns and themes related to motivations and barriers impelling safe riding behaviors among boda-boda riders.

## 4. Results

### 4.1. Quantitative Findings

#### 4.1.1. Characteristics of the study participants

A total of 424 boda-boda riders participated in the study. The mean age of respondents was 29.56 ± 5.71 years, indicating that most riders were young adults. Most riders had attained secondary education (150, 35.4%), suggesting that many riders had basic formal education. Meanwhile, (137, 32.3%) had primary or no formal education, implying that a notable proportion may have limited formal safety training exposure. Most respondents were married (213, 50.2%), indicating that many riders had family responsibilities. Most respondents were Christians (338, 79.7%), reflecting the dominant religion in the study area. Table 1 below shows the details of the participant characteristics.

**Table 1:**
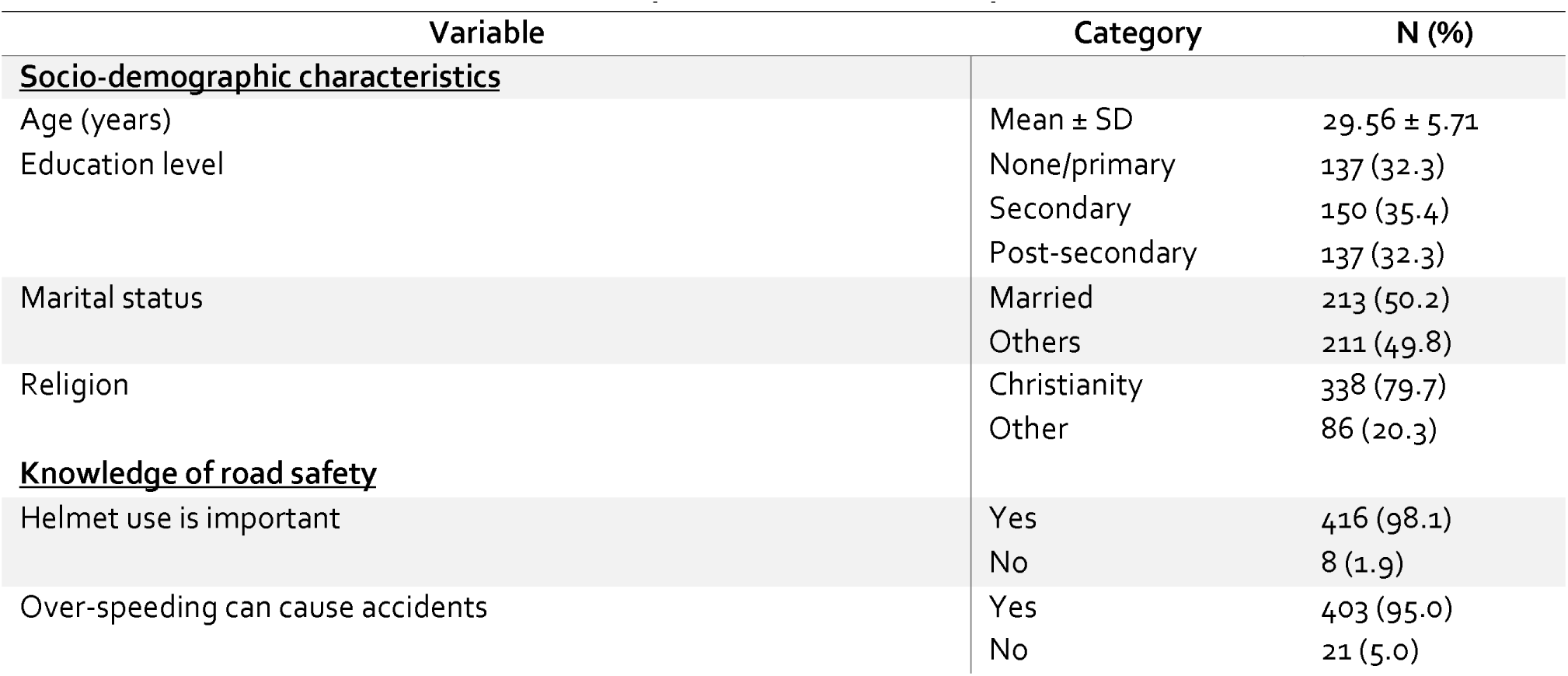
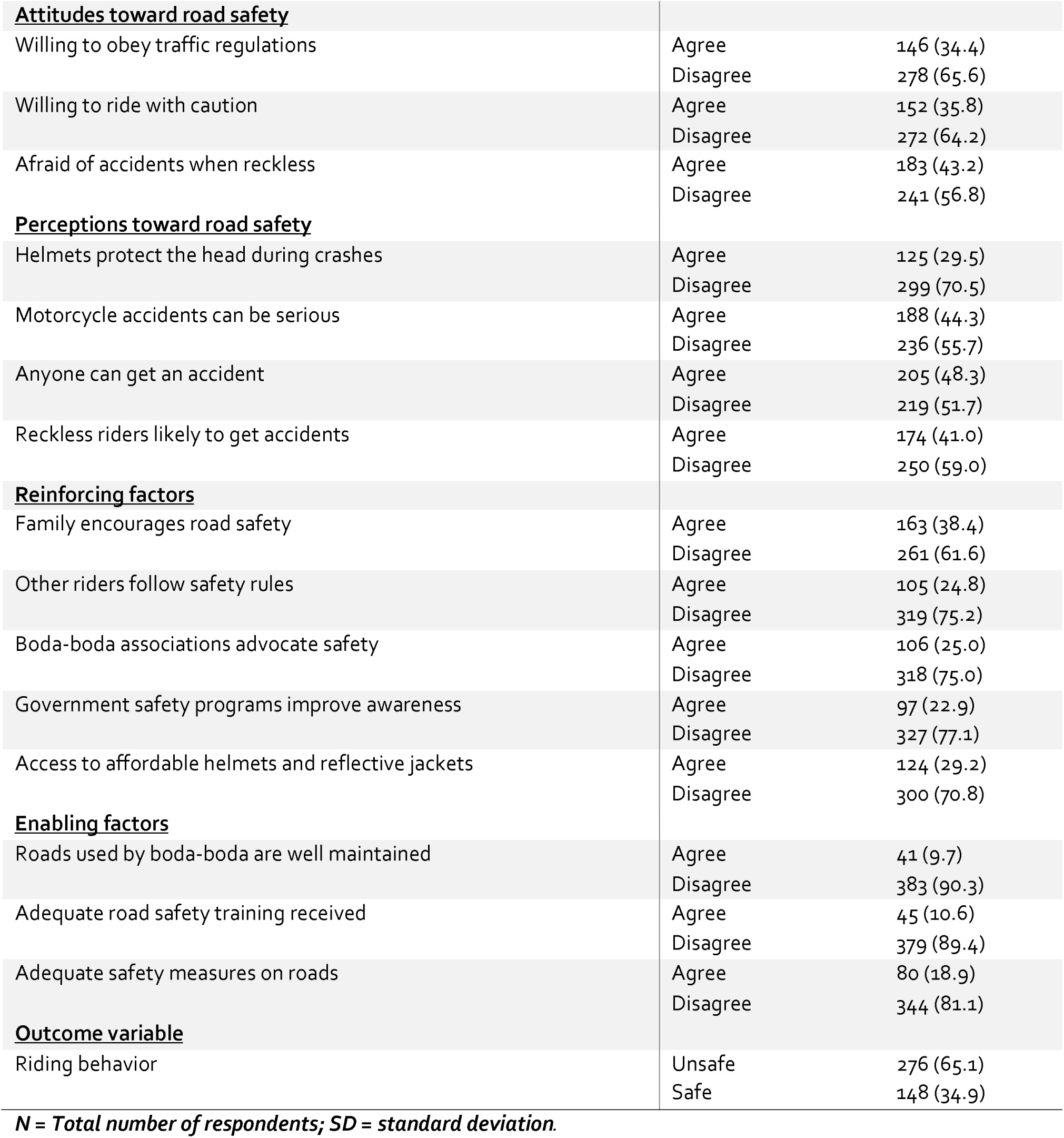
Patient characteristics and descriptive statistics of study variables (N = 424)

| Variable | Category | N (%) |
| --- | --- | --- |
| **<u>Socio-demographic characteristics</u>** |  |  |
| Age (years) | Mean ± SD | $29.56 \pm 5.71$ |
| Education level | None/primary | 137 (32.3) |
|  | Secondary | 150 (35.4) |
|  | Post-secondary | 137 (32.3) |
| Marital status | Married | 213 (50.2) |
|  | Others | 211 (49.8) |
| Religion | Christianity | 338 (79.7) |
|  | Other | 86 (20.3) |
| **<u>Knowledge of road safety</u>** |  |  |
| Helmet use is important | Yes | 416 (98.1) |
|  | No | 8 (1.9) |
| Over-speeding can cause accidents | Yes | 403 (95.0) |
|  | No | 21 (5.0) |
| **<u>Attitudes toward road safety</u>** |  |  |
| Willing to obey traffic regulations | Agree | 146 (34.4) |
|  | Disagree | 278 (65.6) |
| Willing to ride with caution | Agree | 152 (35.8) |
|  | Disagree | 272 (64.2) |
| Afraid of accidents when reckless | Agree | 183 (43.2) |
|  | Disagree | 241 (56.8) |
| **<u>Perceptions toward road safety</u>** |  |  |
| Helmets protect the head during crashes | Agree | 125 (29.5) |
|  | Disagree | 299 (70.5) |
| Motorcycle accidents can be serious | Agree | 188 (44.3) |
|  | Disagree | 236 (55.7) |
| Anyone can get an accident | Agree | 205 (48.3) |
|  | Disagree | 219 (51.7) |
| Reckless riders likely to get accidents | Agree | 174 (41.0) |
|  | Disagree | 250 (59.0) |
| **<u>Reinforcing factors</u>** |  |  |
| Family encourages road safety | Agree | 163 (38.4) |
|  | Disagree | 261 (61.6) |
| Other riders follow safety rules | Agree | 105 (24.8) |
|  | Disagree | 319 (75.2) |
| Boda-boda associations advocate safety | Agree | 106 (25.0) |
|  | Disagree | 318 (75.0) |
| Government safety programs improve awareness | Agree | 97 (22.9) |
|  | Disagree | 327 (77.1) |
| Access to affordable helmets and reflective jackets | Agree | 124 (29.2) |
|  | Disagree | 300 (70.8) |
| **<u>Enabling factors</u>** |  |  |
| Roads used by boda-boda are well maintained | Agree | 41 (9.7) |
|  | Disagree | 383 (90.3) |
| Adequate road safety training received | Agree | 45 (10.6) |
|  | Disagree | 379 (89.4) |
| Adequate safety measures on roads | Agree | 80 (18.9) |
|  | Disagree | 344 (81.1) |
| **<u>Outcome variable</u>** |  |  |
| Riding behavior | Unsafe | 276 (65.1) |
|  | Safe | 148 (34.9) |

*N* = Total number of respondents; *SD* = standard deviation.

Regarding knowledge of road safety, 416 (98.1%) acknowledged the importance of helmet use, indicating very high awareness of helmet safety. Similarly, (403, 95.0%) recognized that over-speeding can lead to accidents, showing that most riders understood key crash risk factors. However, (278, 65.6%) reported unwillingness to obey all traffic regulations, suggesting a deficiency in the knowledge and compliance of riders. Likewise, (272, 64.2%) were not consistently willing to ride with caution, indicating risk-prone attitudes among many riders. Environmental challenges were also reported, with 383 (90.3%) indicating that roads commonly used by boda-boda riders were not well-maintained, suggesting poor road infrastructure. Additionally, (379, 89.4%) reported not receiving adequate road safety training, indicating a lack of formal rider training. Overall, (276, 65.1%) of riders exhibited unsafe riding behaviors, suggesting that unsafe practices were common among most riders.

#### 4.1.2. Bivariate analysis of study variables

In Table 2, bivariate analysis was conducted to examine factors associated with safe riding behavior among boda-boda riders. The mean age of riders practicing unsafe riding behavior (29.74 ± 5.55 years) was comparable to that of riders practicing safe riding behavior (29.23 ± 5.99 years), with no statistically significant difference (t = 0.88, p = 0.381). Among socio-demographic characteristics, marital status (χ² = 4.447, p = 0.035) and religion (χ² = 9.273, p = 0.002) were significantly associated with riding behavior, while education level was not (p = 0.111). Regarding attitudes, willingness to obey traffic regulations was significantly associated with riding behavior (χ² = 8.963, p = 0.003). Other variables, including education level (p = 0.111), anyone can get an accident (p = 0.085), family encouragement for road safety (p = 0.062), access to affordable helmets and reflective jackets (p = 0.064), other riders following safety rules (p = 0.134), and boda-boda associations advocating safety (p = 0.239) also showed potential associations with riding behavior at p ≤ 0.25 and were therefore considered for inclusion in the multivariable logistic regression model.

**Table 2:** Table: Bivariate analysis of factors associated with safe riding behavior among boda-boda riders (N = 424)

| Variable | Category | Unsafe riding behavior n (%) | Safe riding behavior n (%) | Test/Chi-sq statistic | P-value |
| --- | --- | --- | --- | --- | --- |
| **<u>Socio-demographic characteristics</u>** |  |  |  |  |  |
| *Age (years) | Mean ± SD | 29.74 ± 5.55 | 29.23 ± 5.99 | t = 0.88 | 0.381 |
| Education level | None/primary | 80 (58.4) | 57 (41.6) | X<sup>2</sup> = 4.402 | **0.111** |
|  | Secondary | 105 (70.0) | 45 (30.0) |  |  |
|  | Post-secondary | 91 (66.4) | 46 (33.6) |  |  |
| Marital status | Others | 127 (60.2) | 84 (39.8) | X<sup>2</sup> = 4.447 | **0.035** |
|  | Married | 149 (70.0) | 64 (30.0) |  |  |
| Religion | Other | 68 (79.1) | 18 (20.9) | X<sup>2</sup> = 9.273 | **0.002** |
|  | Christianity | 208 (61.5) | 130 (38.5) |  |  |
| **<u>Knowledge of road safety</u>** |  |  |  |  |  |
| Helmet use is important | No | 5 (62.5) | 3 (37.5) | X<sup>2</sup> = 0.024 | 0.876 |
|  | Yes | 271 (65.1) | 145 (34.9) |  |  |
| Over-speeding can cause accidents | No | 13 (61.9) | 8 (38.1) | X<sup>2</sup> = 0.099 | 0.753 |
|  | Yes | 263 (65.3) | 140 (34.7) |  |  |
| **<u>Attitudes toward road safety</u>** |  |  |  |  |  |
| Willing to obey traffic regulations | Disagree | 167 (60.1) | 111 (39.9) | X<sup>2</sup> = 8.963 | **0.003** |
|  | Agree | 109 (74.7) | 37 (25.3) |  |  |
| Willing to ride with caution | Disagree | 166 (61.0) | 106 (39.0) | — | — |
|  | Agree | 110 (72.4) | 42 (27.6) |  |  |
| Afraid of accidents when reckless | Disagree | 154 (63.9) | 87 (36.1) | X<sup>2</sup> = 0.350 | 0.554 |
|  | Agree | 122 (66.7) | 61 (33.3) |  |  |
| **<u>Perceptions toward road safety</u>** |  |  |  |  |  |
| Helmets protect the head during crashes | Disagree | 194 (64.9) | 105 (35.1) | X<sup>2</sup> = 0.020 | 0.888 |
|  | Agree | 82 (65.6) | 43 (34.4) |  |  |
| Motorcycle accidents can be serious | Disagree | 153 (64.8) | 83 (35.2) | X<sup>2</sup> = 0.016 | 0.898 |
|  | Agree | 123 (65.4) | 65 (34.6) |  |  |
| Anyone can get an accident | Disagree | 151 (68.9) | 68 (31.1) | X<sup>2</sup> = 2.963 | **0.085** |
|  | Agree | 125 (61.0) | 80 (39.0) |  |  |
| Reckless riders likely to get accidents | Disagree | 168 (67.2) | 82 (32.8) | X<sup>2</sup> = 1.189 | 0.276 |
|  | Agree | 108 (62.1) | 66 (37.9) |  |  |

| Reinforcing factors |  |  |  |  |  |
| --- | --- | --- | --- | --- | --- |
| Family encourages road safety | Disagree | 161 (61.7) | 100 (38.3) | X<sup>2</sup> = 3.471 | 0.062 |
|  | Agree | 115 (70.6) | 48 (29.4) |  |  |
| Other riders follow safety rules | Disagree | 214 (67.1%) | 105 (32.9%) | X<sup>2</sup> = 2.246 | 0.134 |
|  | Agree | 62 (59.0%) | 43 (41.0%) |  |  |
| Boda-boda associations advocate safety | Disagree | 212 (66.7%) | 106 (33.3%) | X<sup>2</sup> = 1.384 | 0.239 |
|  | Agree | 64 (60.4%) | 42 (39.6%) |  |  |
| Government safety programs improve awareness | Disagree | 215 (65.7%) | 112 (34.3%) | X<sup>2</sup> = 0.270 | 0.603 |
|  | Agree | 61 (62.9%) | 36 (37.1%) |  |  |
| Access to affordable helmets and reflective jackets | Disagree | 187 (62.3%) | 113 (37.7%) | X<sup>2</sup> = 3.442 | 0.064 |
|  | Agree | 89 (71.8%) | 35 (28.2%) |  |  |
| Enabling factors |  |  |  |  |  |
| Roads used by boda-boda are well maintained | Disagree | 246 (64.2%) | 137 (35.8%) | X<sup>2</sup> = 1.303 | 0.254 |
|  | Agree | 30 (73.2%) | 11 (26.8%) |  |  |
| Adequate road safety training received | Disagree | 248 (65.4%) | 131 (34.6%) | X<sup>2</sup> = 0.183 | 0.669 |
|  | Agree | 28 (62.2%) | 17 (37.8%) |  |  |
| Adequate safety measures on roads | Disagree | 225 (65.4%) | 119 (34.6%) | X<sup>2</sup> = 0.078 | 0.779 |
|  | Agree | 51 (63.8%) | 29 (36.3%) |  |  |

$\chi^2$ = Chi-square test; $t$ = independent samples $t$ -test; $SD$ = standard deviation. Statistically significant results at $p \leq 0.25$

#### 4.1.3. Multivariate logistic regression analysis to determine explanatory variables with predictive value for safe riding behaviors

A multivariate logistic regression analysis was conducted to identify factors associated with safe riding behaviors among boda-boda riders. The overall model was statistically significant (χ² (12) = 48.99, p < 0.001), explaining between 10.9% (Cox & Snell R²) and 15.0% (Nagelkerke R²) of the variance in safe riding behaviors. The model correctly classified 68.4% of cases.

Education level was significantly associated with safe riding behaviors. Riders with secondary education (AOR = 0.50, 95% CI: 0.30–0.85, p = 0.011) and post-secondary education (AOR = 0.57, 95% CI: 0.33–0.98, p = 0.041) had lower odds of safe riding behaviors compared to those with none or primary education. Married riders were also less likely to exhibit safe riding behaviors than unmarried riders (AOR = 0.56, 95% CI: 0.34–0.91, p = 0.020).

Riders who identified as Christians had higher odds of safe riding behaviors compared to riders of other religions (AOR = 2.98, 95% CI: 1.63–5.47, p < 0.001). In addition, riders who agreed that roads commonly used by boda-boda are well maintained were more likely to demonstrate safe riding behaviors (AOR = 1.65, 95% CI: 1.07–2.55, p = 0.025). Similarly, riders who reported that boda-boda associations advocate for safety practices had higher odds of safe riding behaviors (AOR = 1.76, 95% CI: 1.03–3.01, p = 0.038).

**Table 3:**
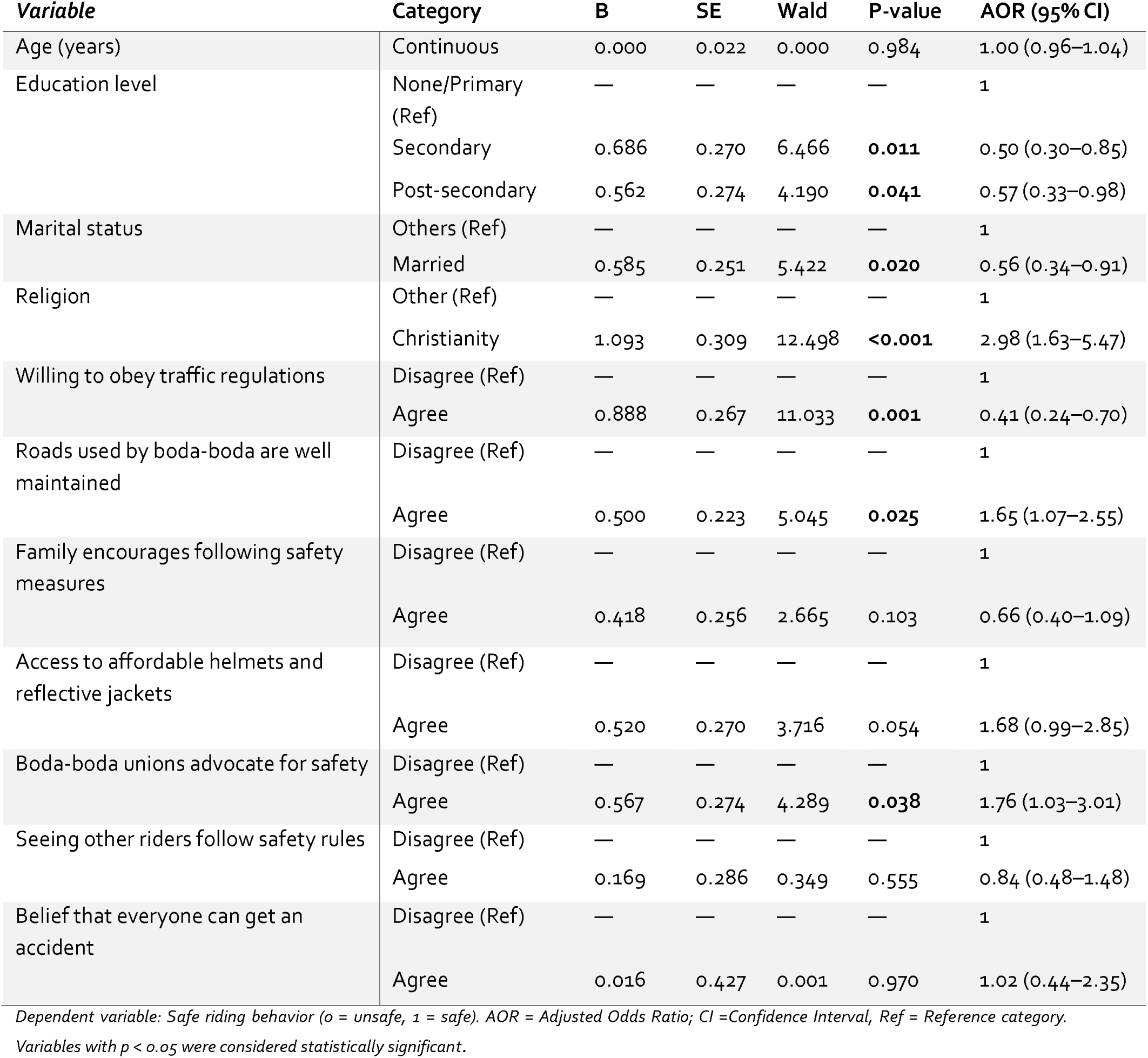
Table: Multivariate analysis of factors associated with safe riding behavior among boda-boda riders (N = 424)

| *Variable* | *Category* | *B* | *SE* | *Wald* | *P-value* | *AOR (95% CI)* |
| --- | --- | --- | --- | --- | --- | --- |
| Age (years) | Continuous | 0.000 | 0.022 | 0.000 | 0.984 | 1.00 (0.96–1.04) |
| Education level | None/Primary (Ref) | — | — | — | — | 1 |
|  | Secondary | 0.686 | 0.270 | 6.466 | **0.011** | 0.50 (0.30–0.85) |
|  | Post-secondary | 0.562 | 0.274 | 4.190 | **0.041** | 0.57 (0.33–0.98) |
| Marital status | Others (Ref) | — | — | — | — | 1 |
|  | Married | 0.585 | 0.251 | 5.422 | **0.020** | 0.56 (0.34–0.91) |
| Religion | Other (Ref) | — | — | — | — | 1 |
|  | Christianity | 1.093 | 0.309 | 12.498 | **<0.001** | 2.98 (1.63–5.47) |
| Willing to obey traffic regulations | Disagree (Ref) | — | — | — | — | 1 |
|  | Agree | 0.888 | 0.267 | 11.033 | **0.001** | 0.41 (0.24–0.70) |
| Roads used by boda-boda are well maintained | Disagree (Ref) | — | — | — | — | 1 |
|  | Agree | 0.500 | 0.223 | 5.045 | **0.025** | 1.65 (1.07–2.55) |
| Family encourages following safety measures | Disagree (Ref) | — | — | — | — | 1 |
|  | Agree | 0.418 | 0.256 | 2.665 | 0.103 | 0.66 (0.40–1.09) |
| Access to affordable helmets and reflective jackets | Disagree (Ref) | — | — | — | — | 1 |
|  | Agree | 0.520 | 0.270 | 3.716 | 0.054 | 1.68 (0.99–2.85) |
| Boda-boda unions advocate for safety | Disagree (Ref) | — | — | — | — | 1 |
|  | Agree | 0.567 | 0.274 | 4.289 | **0.038** | 1.76 (1.03–3.01) |
| Seeing other riders follow safety rules | Disagree (Ref) | — | — | — | — | 1 |
|  | Agree | 0.169 | 0.286 | 0.349 | 0.555 | 0.84 (0.48–1.48) |
| Belief that everyone can get an accident | Disagree (Ref) | — | — | — | — | 1 |
|  | Agree | 0.016 | 0.427 | 0.001 | 0.970 | 1.02 (0.44–2.35) |

*Dependent variable: Safe riding behavior (0 = unsafe, 1 = safe). AOR = Adjusted Odds Ratio; CI = Confidence Interval, Ref = Reference category.*
*Variables with $p < 0.05$ were considered statistically significant.*

### 4.2. Qualitative Findings

Ten passenger interviews (female = 6; male = 4) were thematically analyzed. Five themes emerged: safety concerns, helmet use, passenger responsibility, road infrastructure conditions, and regulation gaps.

#### 4.2.1. Safety concerns

Reported safety ratings ranged from 2/10–8/10. Three passengers rated safety 7/10 (often when using trusted riders or during off-peak travel); two rated 4/10, citing reckless riding and overspeeding; one each rated 8/10, 6/10, and 2/10; two provided qualitative comments without a score. Across interviews, over-speeding (6/10 respondents) and reckless maneuvers/ignoring signals (5/10) predominated; isolated mentions included mechanical faults and limited visible enforcement. Illustrative comments included,

“It depends on the rider… I can give it 7/10” and “It should be 4/10 because they ride recklessly… most are not trained and don’t follow traffic rules.”

#### 4.2.2. Helmet use

7/10 passengers reported not wearing helmets, commonly because riders did not provide one, quality was poor (cracked/dirty/missing liners), or due to comfort/hairstyle concerns. Perceived condition was mixed: clearly poor (3), acceptable (3), mixed (2), or not assessed (2). Two passengers noted better provision/condition when using organized platforms (e.g., SafeBoda, Faras). Even when available, low trust in helmet quality discouraged use.

#### 4.2.3. Passenger responsibility

Passengers described self-protective strategies—refusing rides without helmets, instructing riders to slow down, choosing known riders—yet acknowledged inconsistent self-enforcement. One summarized, “My safety lies in my hands and the boda-boda guy.” Riders’ responsiveness varied; some adjusted their behavior when cautioned, others prioritized speed/income.

#### 4.2.4. Road infrastructure conditions

Perceptions were negative (4) or mixed (4) for many, citing potholes, unmarked bumps, poor drainage, dust/mud, and reduced visibility, with 2 reporting generally good conditions when using trusted routes/riders. These hazards were seen to directly elevate risk for motorcycles.

#### 4.2.5. Regulation gaps

Participants perceived weak enforcement of existing laws (helmet mandates, overloading bans) and called for mandatory licensing/training, periodic safety checks, penalties for non-compliance, and even age-related riding limits. As one noted, “A lot of rules are put on drivers… none are put on the [boda-boda] riders.”

## 5. Discussion

This study examined factors associated with boda-boda rider and passenger safety in Kampala Central Division using a mixed-methods approach guided by the PRECEDE model. The findings indicate that while most riders possess adequate knowledge of safety rules, unsafe practices such as over-speeding, inconsistent helmet use, and neglect of mechanical checks remain widespread. The results demonstrate that attitudinal, social, and structural determinants may play a greater role in shaping safety behavior than demographic or educational characteristics, underscoring the multidimensional nature of boda-boda safety in urban Uganda.

On predisposing factors, education level, specifically secondary and post-secondary, was significant for married Christians. Predisposing factors—including education level, marital status, religion, and attitudes toward obeying traffic regulations—were significantly associated with safe riding behavior in this study. Riders with secondary or post-secondary education and those who were married were less likely to demonstrate safe riding behaviors compared with their counterparts. In contrast, riders identifying as Christians had higher odds of safe riding behaviors. Additionally, willingness to obey traffic regulations emerged as a strong predictor of safer practices.

However, the observed relationship between education and safety behavior in this study contrasts with findings from Nigeria, which reported that riders with higher levels of formal education were more likely to comply with traffic regulations and demonstrate safer riding behaviors ^25,26^. Similarly, a study in Indonesia found that education was positively associated with knowledge and adherence to motorcycle safety practices.

Boda-boda unions’ advocacy on road safety rules was the only significant reinforcing factor. Riders who perceived that their associations actively promoted safety practices were more likely to report safe riding behaviors. This highlights the importance of peer influence, institutional support, and professional regulation within the informal transport sector.

These findings are consistent with studies conducted in Uganda and Tanzania, which have shown that organized motorcycle taxi associations play a critical role in promoting road safety through training, peer monitoring, and enforcement of operational standards ^27,28^. For instance, research on the “SafeBoda” association in Uganda found that its structured training and “sense of community” led to significantly safer riding behaviors compared to regular riders ^29^. Furthermore, a study involving both Uganda and Tanzania demonstrated that such programs can lead to a 43% lower risk of road traffic crashes due to improved knowledge of traffic regulations and safety compliance ^30^. Similarly, motorcycle taxi unions in Tanzania have been recognized for their contributions to improving rider safety through facilitated training programs ^28^.

Conversely, other studies have reported weaker or even negative associations between peer networks and safety behavior. Research done in Bangladesh utilized factor analysis to identify that peer-influenced environments often contribute to safety violations, stunts, and speed violations among young riders ^31^. This is compounded by the fact that over one-fourth of riders in the region admit to regular “ill practices” or misbehavior while riding ^31^. Similarly, a study conducted in Vietnam reported that a rider’s attitude and risk perception, often shaped by social and contextual factors, play a significant role in increasing risky driving behavior rather than reducing it ^32^.

Attitude emerged as a key predictor of safe riding behavior, aligning with evidence from Ghana, Indonesia, and Kenya, where positive attitudes toward safety correlated strongly with compliance with road regulations^33,34^. Although knowledge of safety rules was nearly universal among riders, this did not translate into consistent behavioral adherence, suggesting the well-documented gap between knowledge and practice^35^.

Road conditions were the only strong enabling determinant of safe riding. Poor road surfaces, inadequate signage, and a lack of motorcycle lanes contribute to instability, crashes, and reduced compliance with traffic rules^36 37^. The study found that riders operating on better-maintained routes were more likely to exhibit safer practices, corroborating earlier research in sub-Saharan Africa linking infrastructure quality to road traffic injury prevalence^38^. This result emphasizes that investment in infrastructure is integral to both prevention and mitigation of road traffic injuries.

Despite high awareness of traffic laws, enforcement remains weak. Both riders and passengers described lax application of helmet mandates and overloading restrictions. These findings mirror those of Tumwebaze (2019)^2 6^ and Upano (2021)^6^, who identified weak institutional enforcement as a major barrier to sustainable road safety improvements in Uganda. Riders’ perception that enforcement is inconsistent or easily avoidable undermines compliance and reinforces a culture of impunity. Programs such as SafeBoda have shown promise in institutionalizing enforcement through digital accountability mechanisms and structured training^14,15^, but these scales remain limited. Scaling similar models, while ensuring affordability and institutional support from the Ministry of Works and Transport and the Kampala Capital City Authority (KCCA), could bridge this gap.

This study had some limitations. First, reliance on self-reported data may have introduced recall and social desirability bias, leading to potential underreporting of risky behaviors or overstatement of safety compliance. Second, the sample was limited to Kampala Central Division, where urban dynamics, traffic congestion, and exposure to enforcement differ from those in peri-urban or rural areas, thereby restricting generalizability to other settings.

## 6. Conclusions

In conclusion, boda-boda safety in Kampala is influenced less by demographic characteristics and more by behavioral, social, and infrastructural determinants. Positive attitudes toward safety, supportive family norms, well-maintained roads, and accessible emergency services were associated with safer riding behaviors, while persistent challenges, including inconsistent helmet use, reckless riding, and weak law enforcement, undermine progress. Sustainable improvements will require multisectoral interventions that combine education, enforcement, community engagement, and infrastructural investment. Integrating safety promotion into boda-boda association programs, expanding subsidized access to helmets, improving road maintenance, and strengthening emergency response networks could yield substantial reductions in injury and mortality. Recognizing boda-boda transport as an occupational health priority is essential to advancing urban road safety and protecting both riders and passengers.

## 7. List of abbreviations

AOR: Adjusted Odds Ratio
CI: Confidence Interval
COVID-19: Coronavirus Disease 2019
do: Degrees of Freedom
GLM: Generalized Linear Model
IDI: In-Depth Interview
KCCA: Kampala Capital City Authority
KMO: Kaiser–Meyer–Olkin (measure of sampling adequacy)
LMICs: Low- and Middle-Income Countries
Mowat: Ministry of Works and Transport
MoH: Ministry of Health (Uganda)
OR: Odds Ratio
PCA: Principal Component Analysis
PRECEDE: Predisposing, Reinforcing, and Enabling Constructs in Educational Diagnosis and Evaluation (Model)
REC: Research Ethics Committee
RTI: Road Traffic Injury
SD: Standard Deviation
SE: Standard Error
SPSS: Statistical Package for the Social Sciences
SBI: Safety Behavior Index
WHO: World Health Organization

## 8. Declarations

### 8.1. Ethics approval and consent to participate

Ethical approval for this study was obtained from the Research Ethics Committee (REC) of the Kabale University School of Postgraduate Studies and Research (Approval reference: KABREC-2024-327). Permission to conduct the study was also granted by the Kampala Capital City Authority (KCCA), Department of Public Health and Environment. All participants were informed about the purpose of the study, their role, and their rights, including the right to decline participation or withdraw at any stage without penalty. Written informed consent was obtained from both boda-boda riders and passengers before data collection. Confidentiality and data security were assured, and all information collected was used solely for research purposes. Participants were treated equally, and no personal identifiers were included in the reporting of results. All methods were performed in accordance with the Declaration of Helsinki.

### 8.2. Consent for publication

Not applicable.

### 8.3. Availability of data and materials

All data generated or analysed during this study are included in this published article [and its supplementary information files].

### 8.6. Authors’ contributions

RA and DN conceived the idea. DN analyzed and interpreted the data. RA, DK, DN, TM, SW, EM, JBK, PK, and IM were major contributors in writing and reviewing the manuscript. All authors read and approved the final manuscript.

## 8.7 Acknowledgements

None.

## References

1. WHO, W. H. O. Global status report on road safety 2023. https://www.afro.who.int/sites/default/files/2017-06/vid_global_status_report_en.pdf (2023).

2. Ramdheen, S. & Naicker, B. Evaluating the burden of head injuries on a rural emergency department in South Africa. S Afr Fam Pract (2004) 63, 5327 (2021).

3. Ukachukwu, A.-E. K. et al. Epidemiological Burden of Neurotrauma in Nigeria: A Systematic Review and Pooled Analysis of 45,763 Patients. World Neurosurg 185, e99–e142 (2024).

4. Jean Paul Mvukiyehe. Improving hand hygiene measures in low-resourced intensive care units: experience at the Kigali University Teaching Hospital in Rwanda - PMC. https://pmc.ncbi.nlm.nih.gov/articles/PMC10237047/ (2023).

5. Vaca, S. D. et al. Boda Bodas and Road Traffic Injuries in Uganda: An Overview of Traffic Safety Trends from 2009 to 2017. International Journal of Environmental Research and Public Health 17, 2110 (2020).

6. Godfrey, U. COMMERCIAL CYCLISTS TRAINING AND REDUCTION OF ROAD TRAFFIC ACCIDENTS IN KAMPALA CAPITAL CITY AUTHORITY CENTRAL DIVISION-UGANDA.

7. Tumwebaze, F. Prevalence and factors associated with boda-boda morbidity and mortality at Fort Portal Regional Referral Hospital. http://hdl.handle.net/20.500.12306/4107 (2019).

8. Joana Eva Dodoo. A systematic review of factors leading to occupational injuries and fatalities. https://www.researchgate.net/publication/348306175_A_systematic_review_of_factors_leading_tooccupational_injuries_and_fatalities (2021).

9. Nantongo. A Rapid Assessment of Road Crashes in Uganda: Notes from the Field | Dr. Sulaiman Al Habib Medical Journal. https://link.springer.com/article/10.1007/s44229-022-00018-7 (2022).

10. Atulomah, Y. (PDF) PREDISPOSING-PERSONAL FACTORS ASSOCIATED WITH PASSENGER SAFETY AND SAFE RIDING BEHAVIORS AMONG CYCLISTS (BODA BODA RIDERS) IN KAMPALA. ResearchGate 10.21474/IJAR01/13148 (2021) doi:10.21474/IJAR01/13148.

11. Kasereka, Kamabu Larrey. Predictive models for expansive intracranial hematomas occurrence and surgical evacuation outcomes in traumatic brain injury patients at Mulago Hospital. https://makir.mak.ac.ug/handle/10570/12574 (2023).

12. Sisimwo, P. K. & Onchiri, G. M. Epidemiology of head injuries and helmet use among motorcycle crash injury: a quantitative analysis from a local hospital in Western Kenya. Pan Afr Med J 31, 70 (2018).

13. Konlan, K. D. & Hayford, L. Factors associated with motorcycle-related road traffic crashes in Africa, a Scoping review from 2016 to 2022. BMC Public Health 22, 649 (2022).

14. Doherty, J. Motorcycle taxis, personhood, and the moral landscape of mobility. Geoforum 136, 242–250 (2022).

15. Muni, K. et al. Motorcycle taxi programme increases safe riding behaviours among its drivers in Kampala, Uganda. Injury Prevention 26, 5–10 (2020).

16. Nguyen, T., et al. Injury prevalence and safety habits of boda boda drivers in Moshi, Tanzania: A mixed methods study. PLoS One 13, e0207570 (2018).

17. Nyachieo, G. Levels of Rider Training and Its Influence on Road Safety among Motorcycle (Boda Boda) Riders in Kisumu East Sub-County in Kisumu County, Kenya – Journal of Research Innovation and Implications in Education. https://www.jriiejournal.com/levels-of-rider-training-and-its-influence-on-road-safety-among-motorcycle-boda-boda-riders-in-kisumu-east-sub-county-in-kisumu-county-kenya/ (2021).

18. Yogo, K. O. Training And Regulation Compliance In Motor Cycle Transport Operations In Kisumu City. (University of Nairobi, 2018).

19. AfCAP, A. C. A. P. Bishopetal-AmendTransaidTRL-2019-EnhancingUnderstandingSafeMotorcycleThreeWheelerUse-FinalReport-AfCAP-RAF2114A-190620.pdf. https://www.research4cap.org/wp-content/uploads/ral/Bishopetal-AmendTransaidTRL-2019-EnhancingUnderstandingSafeMotorcycleThreeWheelerUse-FinalReport-AfCAP-RAF2114A-190620.pdf?utm_source=chatgpt.com (2019).

20. Ndagire, M., Kiwanuka, S., Paichadze, N. & Kobusingye, O. Road safety compliance among motorcyclists in Kawempe Division, Kampala, Uganda: a crosssectional study. 10.1080/17457300.2019.1607395 (2019).

21. Tumwesigye, N. M., Atuyambe, L. M. & Kobusingye, O. K. Factors Associated with Injuries among Commercial Motorcyclists: Evidence from a Matched Case Control Study in Kampala City, Uganda | CoLab. https://colab.ws/articles/10.1371%2Fjournal.pone.0148511?utm_source=chatgpt.com (2016).

22. Guest, G., Namey, E. & Chen, M. A simple method to assess and report thematic saturation in qualitative research. PLOS ONE 15, e0232076 (2020).

23. Naeem, M., Ozuem, W., Howell, K. & Ranfagni, S. Demystification and Actualisation of Data Saturation in Qualitative Research Through Thematic Analysis. International Journal of Qualitative Methods 23, 16094069241229777 (2024).

24. Distefano & Yang. Anthony S. DiStefano’s research works | California State University, Fullerton and other places. ResearchGate https://www.researchgate.net/scientific-contributions/Anthony-S-DiStefano-58547968 (2024).

25. Afelumo, O., Abiodun, O. & Sanni, F. Prevalence of Protective Measures and Accident Among Motorcycle Riders with Road Safety Compliance in a Nigerian Semi-Urban Community. Int J Occup Saf Health 11, 129–138 (2021).

26. Awosusi, A. O., Adegboyega, J. A., Adebimpe, W. O. & Osunmakinwa, O. O. Alcohol use and compliance with road safety rules among commercial motorcyclists in Southwest Nigeria. J. Public Health Epidemiol. 13, 38–44 (2021).

27. Mitullah, W. et al. Convergence and divergence of workers’ environment, associations, and access to social protection. in Social Protection and Informal Workers in Sub-Saharan Africa 220–239 (Routledge, London, 2021). doi:10.4324/9781003173694-10.

28. Ntramah, S. et al. Safety, health and environmental impacts of commercial motorcycles in Sub-Saharan African cities. Urban, Planning and Transport Research 11, 2259233 (2023).

29. Mwebesa, M. E., Chou, C.-C., Yoh, K. & Doi, K. A Cross-Sector Framework to Boost the Sustainable Implementation of Integrated Transport and Spatial Strategies to Improve Safety and Mobility of Moto-Taxi Riders. Front. Sustain. Cities 3, 775011 (2021).

30. Gudugbe, S. et al. Approaches to the Effective Prevention of Road Traffic Injuries in Sub-Saharan Africa: A Systematic Review. JSS 11, 323–344 (2023).

31. Hassanzadeh, K., Salarilak, S., Sadeghi-Bazargani, H. & Golestani, M. Motorcyclist risky riding behaviors and its predictors in an Iranian population. J Inj Violence Res 12, 161–170 (2020).

32. Vuong, X. C. et al. Assessing Significant Factors Affecting Risky Riding Behaviors of Vietnamese Motorcyclists Using a Contextual Mediated Model. Journal of Advanced Transportation 2023, 2179828 (2023).

33. Walekhwa. A Rapid Assessment of Road Crashes in Uganda: Notes from the…LJ: Dr. Sulaiman Al Habib Medical Journal. https://journals.lww.com/dshmj/abstract/2022/04040/a_rapid_assessment_of_road_crashes_in_uganda_.2.aspx (2022).

34. Mohammed, A.-R., Yussif, B. G. & Alhassan, M. Road safety attitude and behaviour among motorcycle riders in Ghana: A focus on traffic locus of control and health belief. PLOS ONE 19, e0309117 (2024).

35. Useche, S. A., Montoro, L., Alonso, F. & Tortosa, F. M. Does gender really matter? A structural equation model to explain risky and positive cycling behaviors. Accident Analysis & Prevention 118, 86–95 (2018).

36. Malin, F., Norros, I. & Innamaa, S. Accident risk of road and weather conditions on different road types. Accident Analysis & Prevention 122, 181–188 (2019).

37. Zhang, X., Wen, H., Yamamoto, T. & Zeng, Q. Investigating hazardous factors affecting freeway crash injury severity incorporating real-time weather data: Using a Bayesian multinomial logit model with conditional autoregressive priors. Journal of Safety Research 76, 248–255 (2021).

38. Qiang Zeng. Investigating the Impacts of Real-Time Weather Conditions on Freeway Crash Severity: A Bayesian Spatial Analysis. https://www.mdpi.com/1660-4601/17/8/2768 (2020).

